# Prognostic Value of IL21, CXCL9 and CD1A in Cervical Cancer

**DOI:** 10.64898/2026.01.08.26343702

**Authors:** Yanting Xu, Yunyun Liu, Zhaorong Guo

## Abstract

**Background:** Cervical cancer is one of the most common malignant tumors of the female reproductive system. Existing treatments provide limited benefit for patients with advanced, recurrent or metastatic disease, and reliable prognostic markers are lacking. In this study we integrated multi-omic data from The Cancer Genome Atlas (TCGA) and the Genotype-Tissue Expression (GTEx) database. Protein-coding genes meeting the criteria of an adjusted P value < 0.05 and |log_2_ fold-change| > 5 were screened; 693 genes were identified. We further focused on three genes related to the tumor microenvironment—interleukin 21 (IL21), C-X-C motif chemokine ligand 9 (CXCL9) and cluster of differentiation 1A (CD1A)—and performed differential expression analysis, survival analysis, clinical stage analysis and immune infiltration correlation analysis to clarify their prognostic value and potential mechanisms in cervical cancer.

**Results:** (1) CXCL9 and CD1A were highly expressed in cervical cancer tissues, and all three genes showed high expression across different pathological stages without stage-dependent differences; (2) high expression of IL21, CXCL9 and CD1A improved patient prognosis and was positively associated with overall survival (OS), disease-specific survival (DSS) and progression-free interval (PFI); (3) expression of IL21, CXCL9 and CD1A was closely correlated with infiltration of multiple immune cells: IL21 correlated with total T cells, helper T cells and B cells, CXCL9 correlated with T cells and activated dendritic cells, and CD1A correlated with immature dendritic cells.

**Conclusion:** IL21, CXCL9 and CD1A are potential prognostic biomarkers and key immunomodulatory factors in cervical cancer. This study provides a new direction for immunotherapy and individualized precision treatment of cervical cancer.

## 1. Introduction

Cervical cancer is a common malignancy of the female reproductive system characterized by a high incidence and poor prognosis. Persistent infection with human papillomavirus (HPV) is the principal etiological factor. Current treatment modalities include surgery, radiotherapy, chemotherapy, targeted drugs and immunotherapy; however, therapeutic effects for advanced, metastatic or recurrent disease remain unsatisfactory^[1]^. In recent years, the tumor microenvironment (TME) has received increasing attention because it consists of immune cells, stromal cells and vascular endothelial cells, and the level and functional state of immune cell infiltration are closely related to patient prognosis^[2]^. Interleukin-21 (IL-21) is an immunostimulatory cytokine belonging to the common γ-chain family. It plays a pivotal role. IL-21 is primarily produced by CD4+ T follicular helper and Th17 cells; activation of the IL-21 receptor recruits JAKs and activates STAT3, PI3K and MAPK pathways, promoting the proliferation and differentiation of B, T and NK cells and formation of stem-like memory CD8+ T cells^[3]^. Preclinical and clinical studies have shown that IL-21 has antitumor activity with minimal side effects, making it a promising target for cancer therapy^[4]^.

CXCL9 is a chemokine that binds CXCR3 and recruits cytotoxic lymphocytes, NK cells, NKT cells and macrophages into tumors^[5]^. High CXCL9 expression has been shown to correlate with better survival in cervical cancer patients^[6]^, and engineered cells co-expressing IL-21 and CXCL9 display enhanced expansion and infiltration, underscoring the antitumor effects of these ^[7]^.

CD1A encodes a lipid-antigen–presenting molecule expressed on dendritic cells^[8]^. Earlier studies showed that CD1a+ dendritic cells are less abundant in HPV-associated cervical lesions than in normal epithelium, suggesting that reduced antigen presentation contributes to immune ^[9]^.

Although IL21, CXCL9 and CD1A participate in immune regulation and tumor progression, their prognostic value and immunomodulatory mechanisms in cervical cancer remain unclear. This study systematically evaluates the expression patterns and prognostic significance of IL21, CXCL9 and CD1A using TCGA and GTEx data. It aims to clarify the roles of these genes in regulating the cervical cancer microenvironment, provide theoretical support for developing new immunotherapies, and ultimately improve patient outcomes.

## 2. Materials and Methods

### 2.1 Data sources

Transcriptomic data (RNA-Seq, TPM format) and corresponding clinical information (including TNM stage, survival time and status) were obtained from the Cervical Squamous Cell Carcinoma and Endocervical Adenocarcinoma (CESC) project of TCGA (https://portal.gdc.cancer.gov/). Expression data for 309 tumor samples and three normal tissues were retrieved. After excluding samples with incomplete clinical information and duplicates, 241 tumor samples and three normal tissues were included for subsequent analyses. Differential expression data were supplemented using the TCGA–GTEx dataset from the UCSC XENA database, which comprised 306 cervical cancer tissues and 13 normal tissues.

### 2.2 Target gene screening

Protein-coding genes were screened using the criteria of adjusted P value (Padj) < 0.05 and |log_2_FC| > 5, yielding 693 differentially expressed genes. Prognostic analyses for overall survival (OS), disease-specific survival (DSS) and progression-free interval (PFI) narrowed the candidates to 13 genes significantly associated with prognosis. Among these, three genes—IL21, CXCL9 and CD1A—were closely related to the tumor microenvironment and were selected as the focus of this study.

### 2.3 Differential expression analysis

The ggplot2 package (v3.4.4) was used to visualize the expression differences of IL21, CXCL9 and CD1A between normal and cervical cancer tissues. Statistical analyses were performed using the stats (v4.2.1) and car (v3.1-0) packages.

### 2.4 Survival analysis

Receiver operating characteristic (ROC) curves were generated to assess the diagnostic efficacy of IL21, CXCL9 and CD1A for cervical cancer prognosis. The pROC package (v1.18.0) was used for ROC analysis and ggplot2 for visualization. The area under the curve (AUC) was used to evaluate predictive performance. Kaplan–Meier survival analyses were performed with the online tools and packages survival (v3.3.1), ggplot2 (v3.4.4) and survminer (v0.4.9). Log-rank tests and Cox regression analyses assessed the influence of gene expression on OS, DSS and PFI. Statistical significance was defined as P < 0.05.

### 2.5 Clinical stage analysis

Tumor samples were stratified into four groups according to clinical stage (I–IV). The Kruskal–Wallis test (for multiple groups) or Wilcoxon rank-sum test (for two groups) evaluated expression differences between normal tissue and different stages, and among stages. The results were visualized using ggplot2, stats and car packages.

### 2.6 Immune infiltration correlation analysis

Immune infiltration analyses assessed correlations between IL21, CXCL9 and CD1A expression and immune cell subsets including activated dendritic cells (aDC), B cells, CD8+ T cells, cytotoxic cells, dendritic cells (DC), eosinophils, immature dendritic cells (iDC), macrophages, mast cells, neutrophils, NK CD56bright cells, NK CD56dim cells, natural killer (NK) cells, plasmacytoid DCs (pDC), T cells, helper T cells, central memory T cells (Tcm), effector memory T cells (Tem), follicular helper T cells (TFH), γδ T cells, Th1 cells, Th17 cells, Th2 cells and regulatory T cells (Treg). The ggplot2 package was used for visualization. All analyses were performed in R (v4.2.1).

### 2.7 Statistical analysis

Wilcoxon rank-sum tests compared gene expression between normal and cancer tissues. Kruskal–Wallis tests with Dunn’s post-hoc tests and Wilcoxon tests explored relationships between gene expression and clinical parameters. Time-dependent ROC curves assessed prognostic diagnostic efficacy. Kaplan–Meier curves, log-rank tests and Cox regression analyses evaluated associations with OS, DSS and PFI. Spearman correlation analyses assessed relationships between gene expression and immune infiltration. P values < 0.05 were considered statistically significant.

## 3. Results

### 3.1 Up-regulation of CXCL9 and CD1A in cervical cancer

CXCL9 and CD1A were significantly up-regulated in cervical cancer tissues compared with normal tissues (P < 0.05). Due to the lack of expression data in normal tissues, IL21 was not compared between normal and cancer tissues.(Figure 1)

**Figure 1.**
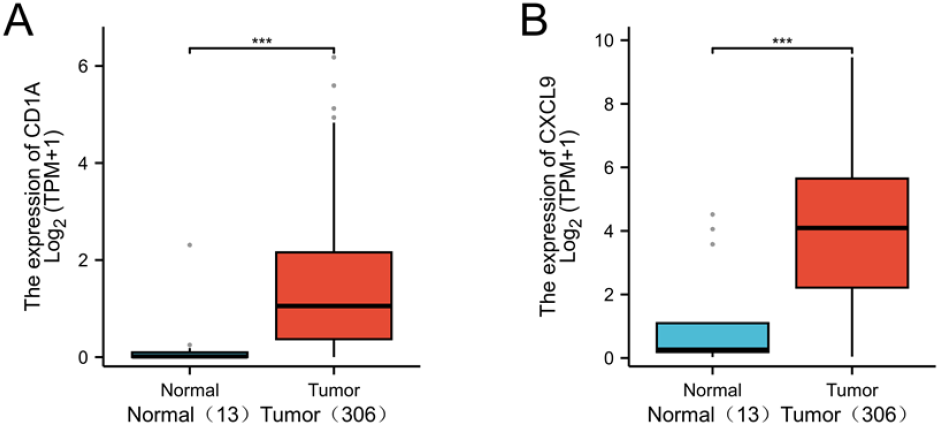
(A) Expression levels of CD1A and (B) CXCL9 in cervical cancer patients and matched normal samples, respectively.P < 0.05.

### 3.2 Relationship between gene expression and clinical stage

Expression of CXCL9 and CD1A in stages I–IV was significantly higher than in normal tissues (P < 0.05). However, no statistically significant differences in IL21, CXCL9 or CD1A expression were observed among the four stages, indicating that these genes are highly expressed from early disease onward without stage-dependent changes.(Figure 2)

**Figure 2.**
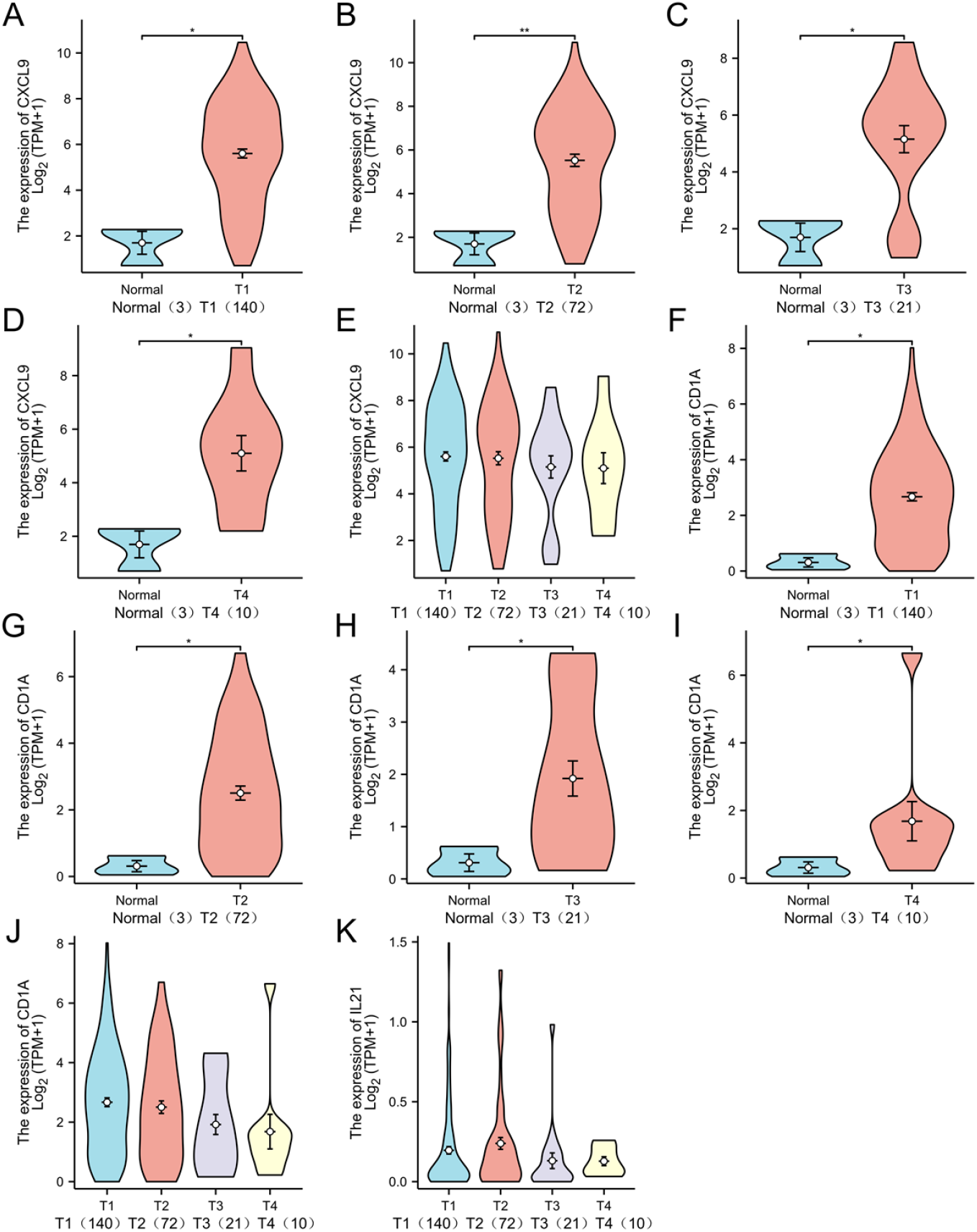
(A-E) Expression of CXCL9 and (F-J) CD1A in cervical cancer at different pathological stages (I, II, III, and IV) and the expression differences among different pathological stages;(A-D),(F-I)P<0.05;(E),(J)P>0.05 (K) Expression differences of IL21 among different pathological stages.P>0.05.

### 3.3 Prognostic predictive value

ROC curve analysis showed that the AUCs for IL21, CXCL9 and CD1A were 0.894, 0.918 and 0.900, respectively, indicating good predictive accuracy. Kaplan–Meier analyses demonstrated that high expression of IL21, CXCL9 and CD1A was associated with significantly improved OS, DSS and PFI (P < 0.05).(Figure 3)

**Figure 3.**
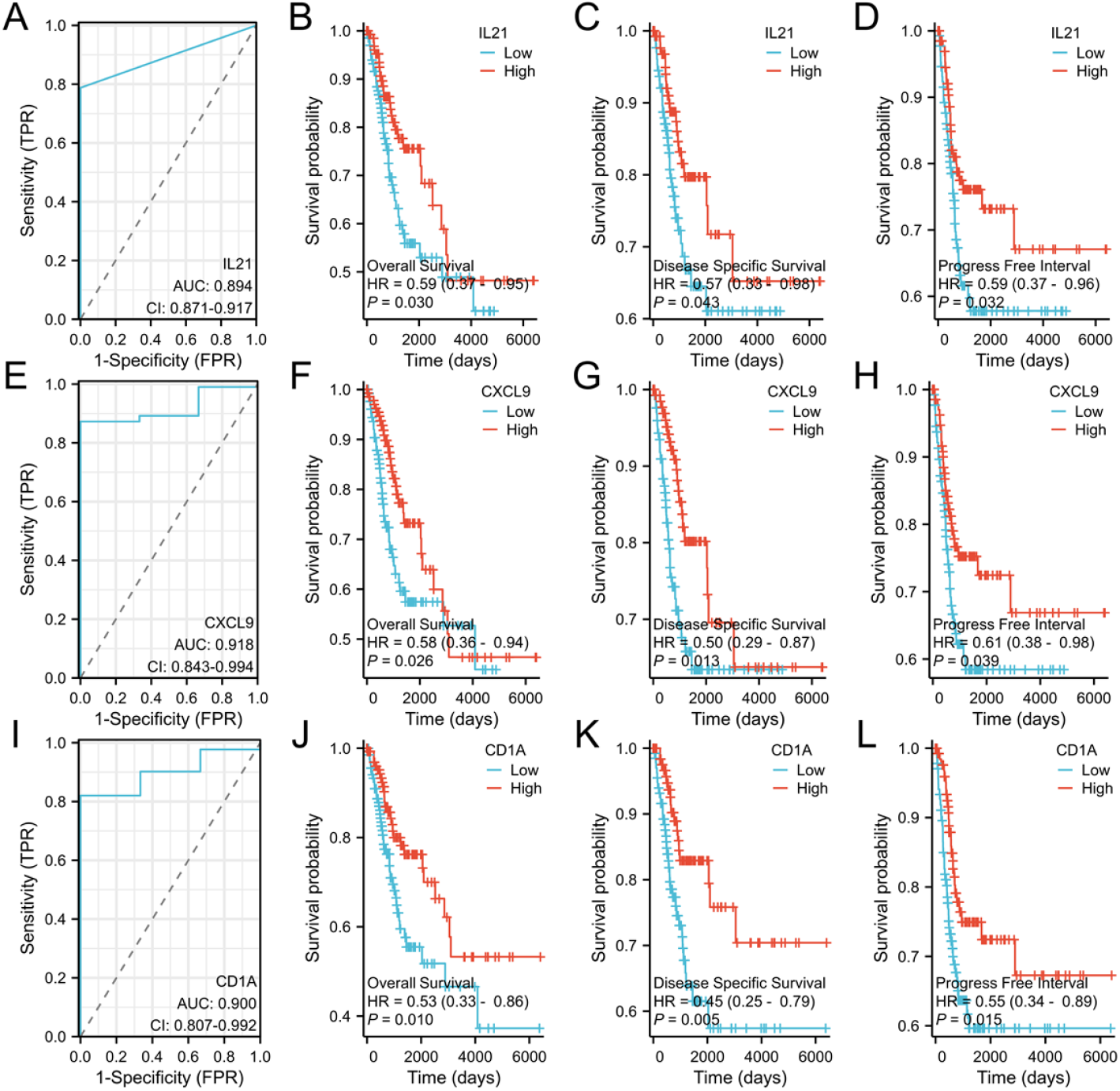
(A, E, I) The receiver operating characteristic (ROC) curve respectively reflects the diagnostic capabilities of IL21, CXCL9, and CD1A for cervical cancer; the Kaplan-Meier (KM) graph shows the effects of IL21, CXCL9, and CD1A on the overall survival (OS) (B, F, J), disease-specific survival (DSS) (C, G, K), and progression-free survival (PFI) (D, H, L) of cervical cancer patients.P<0.05

### 3.4 Immune infiltration correlation

Expression of IL21, CXCL9 and CD1A correlated positively with multiple immune cell subsets (R > 0, P < 0.05) including aDC, B cells, CD8+ T cells, cytotoxic cells, DC, iDC, mast cells, neutrophils, NK CD56dim cells, helper T cells, Tcm, TFH, Th1 cells, Th2 cells and Treg. Notably, IL21 was strongly correlated with total T cells, helper T cells and B cells (correlation coefficient R > 0.6); CXCL9 was strongly correlated with T cells and aDC (R > 0.7); and CD1A was strongly correlated with iDC (R > 0.6).(Figure 4)

**Figure 4.**
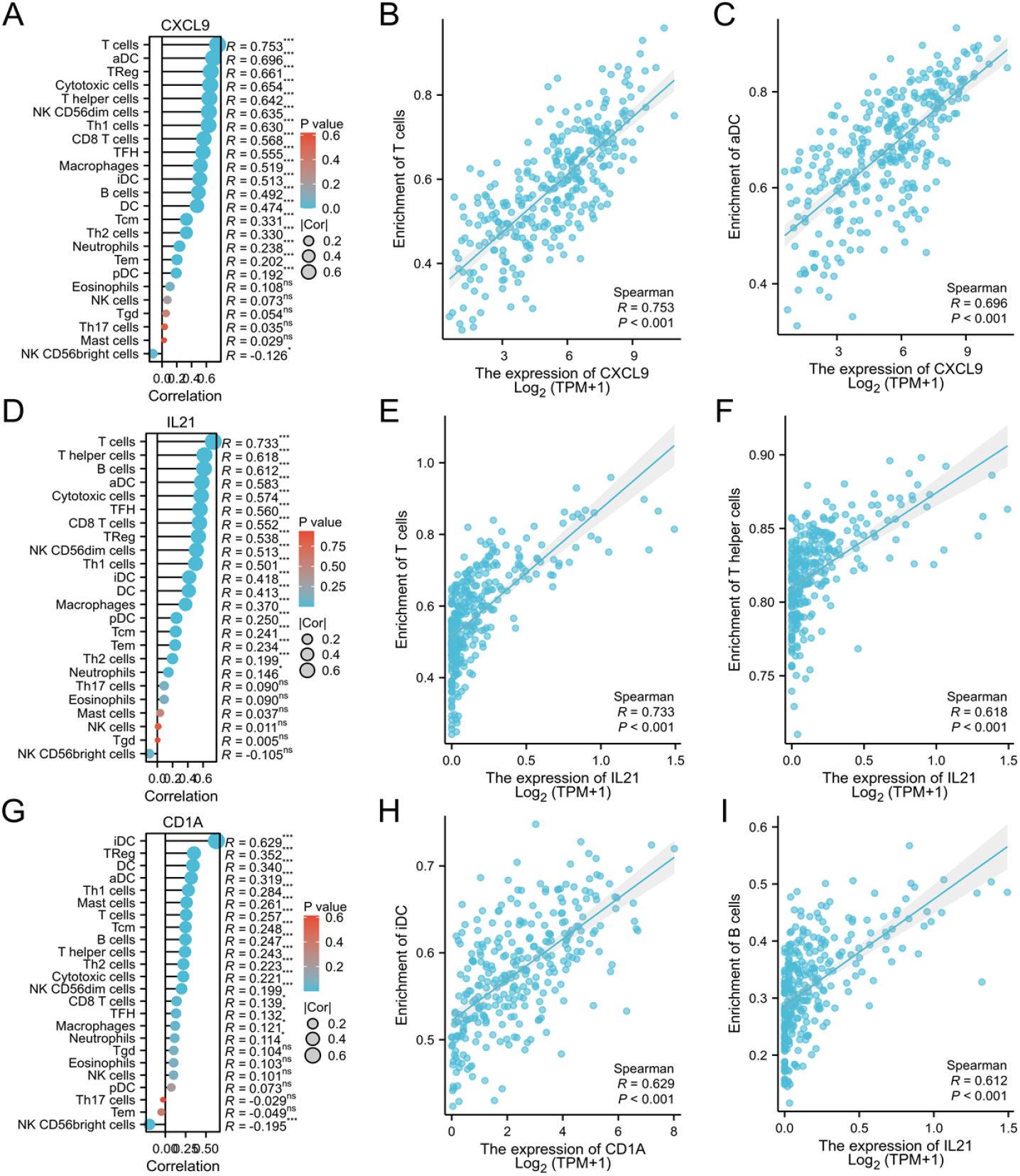
The expression of CXCL9, IL21 and CD1A is correlated with the infiltration of various immune cells (A, D, G); CXCL9 is significantly correlated with T cells and aDC (R > 0.7) (B, C); IL21 is significantly correlated with T cells, helper T cells, and B cells (R > 0.6) (E, F, I); CD1A is significantly correlated with iDC (R > 0.6) (H).P<0.05

## 4. Discussion

Cervical cancer remains one of the most prevalent gynecological malignancies and is dominated by squamous cell carcinoma and adenocarcinoma. Its pathogenesis involves viral infection, immune evasion and the tumor microenvironment. Although surgery, radiotherapy and chemotherapy can benefit some patients, outcomes for advanced, metastatic or recurrent cases are still poor. Therefore, exploring molecular mechanisms and their interactions with the microenvironment is essential for discovering new therapeutic targets and improving prognosis.

Using stringent criteria, this study identified IL21, CXCL9 and CD1A as differentially expressed genes associated with survival in cervical cancer. Differential expression analysis indicated that CXCL9 and CD1A were significantly up-regulated in tumor tissues, whereas IL21 lacked comparable normal data. Expression of all three genes was consistently high across stages I–IV, suggesting early involvement in tumorigenesis rather than stage-dependent increases. This contrasts with the conventional view that oncogenes are gradually up-regulated during tumor progression^[10]^. One explanation is that CXCL9 and CD1A participate mainly in initiating immune surveillance, promoting early immune cell infiltration and antigen recognition; during late stages tumors may bypass these mechanisms, resulting in stable expression^[11]^. Clinically, targeting these molecules may be beneficial irrespective of stage.

High expression of IL21, CXCL9 and CD1A was associated with longer OS, DSS and PFI. IL-21 exerts antitumor effects by activating CD8+ T cells and helper T cells and enhancing adaptive immunity^[3]^. CXCL9 recruits cytotoxic lymphocytes, NK cells and macrophages via the CXCL9/CXCR3 axis, improving immune infiltration and survival, and its high expression correlates with favorable prognosis^[5]^. CD1A encodes an antigen-presenting molecule expressed on dendritic cells^[8]^. Reduced numbers of CD1a+ dendritic cells were found in HPV-associated cervical lesions, suggesting that high CD1A expression reflects robust antigen presentation and immune surveillance^[9]^.

Immune infiltration correlation analyses revealed that IL21, CXCL9 and CD1A expression was positively correlated with a broad range of immune cell subsets, with particularly strong associations between IL21 and helper T/B cells, CXCL9 and activated dendritic cells, and CD1A and immature dendritic cells. CD8+ T cells are key effectors in antitumor immunity but may become exhausted under chronic antigen stimulation, leading to reduced proliferation and cytokine secretion^[12]^. B cells not only produce antibodies but also present antigens efficiently, amplifying T-cell responses^[13]^. Dendritic cells are essential for cross-presentation of exogenous antigens and activation of CD8+ T cells^[14]^, while Th1 cells secrete interferon-γ to activate macrophages and CD8+ T cells^[15]^. Our findings suggest that IL21, CXCL9 and CD1A improve prognosis by promoting infiltration and activation of these immune populations.

Given the multifunctional nature of IL-21, targeted delivery to specific immune subsets has been proposed to enhance antitumor efficacy while avoiding inhibitory effects on dendritic cells. CXCL9-driven recruitment of cytotoxic cells may synergize with IL-21-mediated T-cell activation, and CD1A-positive dendritic cells can present lipid antigens to T cells, forming a cooperative network. Agents such as norcantharidin have been shown to modulate immune cell function and remodel the tumor microenvironment^[16]^, providing a rationale for combination therapies. Our study provides a theoretical basis for exploring immunomodulators targeting IL21, CXCL9 and CD1A to improve outcomes in cervical cancer.

## 5. Conclusion

This study identifies IL21, CXCL9 and CD1A as potential prognostic biomarkers and key immunoregulatory factors in cervical cancer. High expression of these genes correlates with improved survival and enhanced immune infiltration. Our findings provide new insights into the tumor microenvironment and offer theoretical support for immunotherapy and individualized precision treatment of cervical cancer. Future work should further investigate agents that modulate these pathways, such as norcantharidin, to translate these discoveries into clinical applications.

## Data Availability

All relevant data are within the manuscript and its Supporting Information files.

https://portal.gdc.cancer.gov/

## Availability of Data and Materials

The datasets generated and/or analysed during the current study are available in the TCGA database (https://portal.gdc.cancer.gov/) and XENA database (https://xenabrowser.net/datapages/?host= https://toil.xenahubs.net)

## Author Contributions

XYT were responsible for the design of the study protocol. GZR and LYY contributed to the writing of the study protocol and assisted in completing statistical analysis and picture drawing. All authors contributed to manuscript revision, read, and approved the submitted version. All authors have participated sufficiently in the work and agreed to be accountable for all aspects of the work.

## Ethics Approval and Consent to Participate

TCGA and GEO databases are publicly available and written informed consent was obtained from the patients prior to data collection.

## Acknowledgment

Not applicable.

## References

[1] Chao X, Song X, Wu H, You Y, Wu M, Li L. Selection of Treatment Regimens for Recurrent Cervical Cancer. Front Oncol. 2021;11:618485. doi:10.3389/fonc.2021.618485

[2] Lee J, Shin S, Kim JH, et al. Uterine Cervical Angioleiomyoma Mimicking Squamous Cell Carcinoma. Diagnostics (Basel). 2023;13(14):2370. doi:10.3390/diagnostics13142370

[3] Topchyan P, Xin G, Chen Y, et al. Harnessing the IL-21-BATF Pathway in the CD8^+^ T Cell Anti-Tumor Response. Cancers (Basel). 2021;13(6). Published 2021 Mar 12. doi:10.3390/cancers13061263

[4] Isvoranu G, Chiritoiu-Butnaru M. Therapeutic potential of interleukin-21 in cancer. Front Immunol. 15:1369743. Published 2024 None. doi:10.3389/fimmu.2024.1369743

[5] Xiong TF, Pan FQ, Liang Q, et al. Prognostic value of the expression of chemokines and their receptors in regional lymph nodes of melanoma patients. J Cell Mol Med. 2020;24(6):3407–3418. doi:10.1111/jcmm.15015

[6] Yang L, Yang Y, Meng M, et al. Identification of prognosis-related genes in the cervical cancer immune microenvironment. Gene. 766:145119. doi:10.1016/j.gene.2020.145119

[7] Chen S, Gong F, Liu S, et al. IL-21- and CXCL9-engineered GPC3-specific CAR-T cells combined with PD-1 blockade enhance cytotoxic activities against hepatocellular carcinoma. Clin Exp Med. 2024;24(1):204. Published 2024 Aug 28. doi:10.1007/s10238-024-01473-2

[8] Gulic T, Laskarin G, Glavan L, Grubić Kezele T, Haller H, Rukavina D. Human Decidual CD1a<sup>+</sup> Dendritic Cells Undergo Functional Maturation Program Mediated by Gp96. Int J Mol Sci. 2023;24(3). Published 2023 Jan 23. doi:10.3390/ijms24032278

[9] Jesus ACC, Meniconi MCG, Galo LK, Duarte MIS, Sotto MN, Pagliari C. Plasmacytoid Dendritic Cells, the Expression of the Stimulator of Interferon Genes Protein (STING) and a Possible Role of Th17 Immune Response in Cervical Lesions Mediated by Human Papillomavirus. Indian J Microbiol. 2023;63(4):588–595. doi:10.1007/s12088-023-01117-1

[10] S D A, Pasumarthi D, Pasha A, et al. Identification of Differentially Expressed Genes in Cervical Cancer Patients by Comparative Transcriptome Analysis. Biomed Res Int. 2021;2021:8810074. doi:10.1155/2021/8810074

[11] Giannos P, Kechagias KS, Bowden S, Tabassum N, Paraskevaidi M, Kyrgiou M. PCNA in Cervical Intraepithelial Neoplasia and Cervical Cancer: An Interaction Network Analysis of Differentially Expressed Genes. Front Oncol. 2021;11:779042. doi:10.3389/fonc.2021.779042

[12] Zhang Q, Zhang W, Lin T, et al. Mass cytometry reveals immune atlas of urothelial carcinoma. BMC Cancer. 2022;22(1):677. doi:10.1186/s12885-022-09788-7

[13] Yarchoan M, Ho WJ, Mohan A, et al. Effects of B cell-activating factor on tumor immunity. JCI Insight. 2020;5(10):e136417. doi:10.1172/jci.insight.136417

[14] MacNabb BW, Tumuluru S, Chen X, et al. Dendritic cells can prime anti-tumor CD8? T cell responses through major histocompatibility complex cross-dressing. Immunity. 2022;55(6):982-997.e8. doi:10.1016/j.immuni.2022.04.016

[15] Ryba-Stanisławowska M. Unraveling Th subsets: insights into their role in immune checkpoint inhibitor therapy. Cell Oncol (Dordr). 2025;48(2):295–312. doi:10.1007/s13402-024-00992-0

[16] Tao Y, Shen L, Wang P. Advances and perspectives on isolation, structural characterization, structure-property relationships, and pharmacological mechanisms of tumor immune glucans: A review. Int J Biol Macromol. 2025;315(Pt 2):144621. doi:10.1016/j.ijbiomac.2025.144621

